# Knowledge, Attitude and Perceptions Regarding Male Infertility among the Medical Students and Healthcare Professionals of Bangladesh

**DOI:** 10.1101/2022.02.18.22271175

**Authors:** Mohammad Azmain Iktidar, A M Khairul Islam, Sreshtha Chowdhury, Simanta Roy, Mahzabeen Islam, Tonmoy Chowdhury, Mustari Nailah Tabassum, Tahsin Sumat Ali, Atandra Akash, Mashrur Ahmed, Faraz Al Zafar, Mohammad Delwer Hossain Hawlader

## Abstract

**Background:** People worldwide have many misconceptions regarding reproductive health and fertility because infertility is still a taboo subject. The general public seek medical and psychological counsel and assistance from health experts about male infertility. Therefore, this study aimed to explore the knowledge, attitude, and behavior regarding male infertility among medical students and health care workers of Bangladesh.

**Method:** This cross-sectional study used quota sampling to assure equal participation from each of the eight divisions of Bangladesh and convenience. 46 structured questions were used to assess respondents’ knowledge, attitudes, and perceptions (34-items for knowledge, 5-items for attitudes, and 7-items for perceptions).Two scoring systems were employed with the mean scores as cut-off value for good knowledge and positive attitude. The mean knowledge and attitude scores were then correlated with sociodemographic factors using chi-square and two-independent sample t-tests. Finally, we performed binary logistic regression to explore predictors of good knowledge and positive attitude.

**Result:** Among the participant, 49.82% did not have a good male infertility knowledge and nearly 60.79% had negative attitudes regarding male infertility. Young (23-26 years) healthcare professionals and medical students were more likely to have good knowledge than others (OR: 1.81; 95% CI: 1.099 to 2.988). Surprisingly, females were more likely to have positive attitude (OR=1.48; 95%CI: 1.002 and 2.19, p=0.049) compared to males. Among all the professions, MBBS doctors were most likely to have good knowledge and positive attitude regarding male infertility. Good knowledge of male infertility predicted positive attitude (OR=1.61; 95% CI: 1.105 and 2.346, p=0.013) and vice versa.

**Conclusion:** Our research found that despite favourable opinions, healthcare professionals and medical students in Bangladesh had inadequate knowledge and negative attitudes regarding male infertility. This emphasizes the need for interdisciplinary training programs, standardization of healthcare worker guidelines, and curricular adjustments for medical students.

## Introduction

Childbirth is seen as a crucial component of human existence, and most men and women take parenting for granted and look forward to it. Infertility is a condition of the reproductive system that affects both men and women at almost the same rate, and it is a common phenomenon. ^1^According to WHO, Infertility is a disorder of the male or female reproductive system characterized by the inability to conceive after 12 months or more of unprotected sexual activity. Around the world, more than 70 million couples of the global population suffer from infertility, and the majority of them are residents of developing countries.^2^ Infertility patterns in poor countries differ significantly from those in developed countries. It may be distressing for couples because societal conventions and religious dictums may associate infertility with personal, interpersonal, emotional, or social failure. In the majority of situations, women suffer the burden of cultural prejudices.^3^

Although it was formerly thought that infertility was primarily due to a female element, it is now widely acknowledged that male factor infertility is just as essential as female factor infertility.^4^ Male infertility accounts for a considerable share of infertility, and reproductive tract infections, STDs, varicocele, diabetes, obesity, cystic fibrosis, hypogonadism are the major causes to be seen. ^5^ Tight undergarments, keeping mobile or laptops near the genitalia, tobacco, and excessive exercise have also been proved to be responsible for decreased sperm concentration in the male population.

The prevalence of male infertility has not been accurately recognized due to patriarchal preferences in many countries.^6^ Due to a lack of understanding of the factors contributing to male infertility, some men may unknowingly engage in activities that impair their capacity to produce biological children.^7^ Certain medical conditions like cancer, genitourinary surgery, prostate surgery, cardiovascular disease can affect the fertility rate to a great extent. So, there is a need to educate health service providers about this issue and improve men’s awareness of avoiding and managing reproductive health issues.

People worldwide have many misconceptions regarding reproductive health and fertility because infertility is still a taboo subject.^8^ Understanding psychological and social problems associated with infertility would benefit from recognizing ideas and attitudes towards male infertility.^9^ Uses of alternative medicines like herbal, spiritual healing, homeopathy etc., are pretty popular in male infertility treatment. In developing countries, people tend to have more affinity towards these than other medical treatments. However, it is much more important to introduce reproductive health through mass media and campaigns in educational institutions. Also, a complex mix of social, cultural, and medical factors must be included in these awareness programs.

Men face a difficult emotional journey after getting diagnosed with infertility disorder. According to research, infertile males have poorer self-esteem, higher anxiety, and more somatic symptoms.^10^ Many studies investigated infertility-related knowledge, behaviors, perceptions, and practices, but only a small amount of information is known for male infertility-related knowledge, attitude and perceptions. So, male infertility is a grave concern in our family and society. The general public seek medical and psychological counsel and assistance from health experts about male infertility. So, this issue should be well-understood by all physicians, medical students, and other health care providers who will become doctors in the future. This study aimed to explore the knowledge, attitude, and perceptions regarding male infertility among medical students and health care workers of Bangladesh.

## Method

### Study participants and study site

This cross-sectional study was done in Bangladesh among 556 medical students and healthcare professionals (Figure 1). The research was completed in two months. The researchers employed an online survey to ensure social distancing and take appropriate precautions throughout the pandemic. Sample were selected quota sampling method to ensure equal representation from each of the eight divisions of Bangladesh and convenience. Participants were included upon meeting following criteria: (1) Bangladeshi resident, (2) health care professional or medical student, and (3) providing informed consent

**Figure 1.**
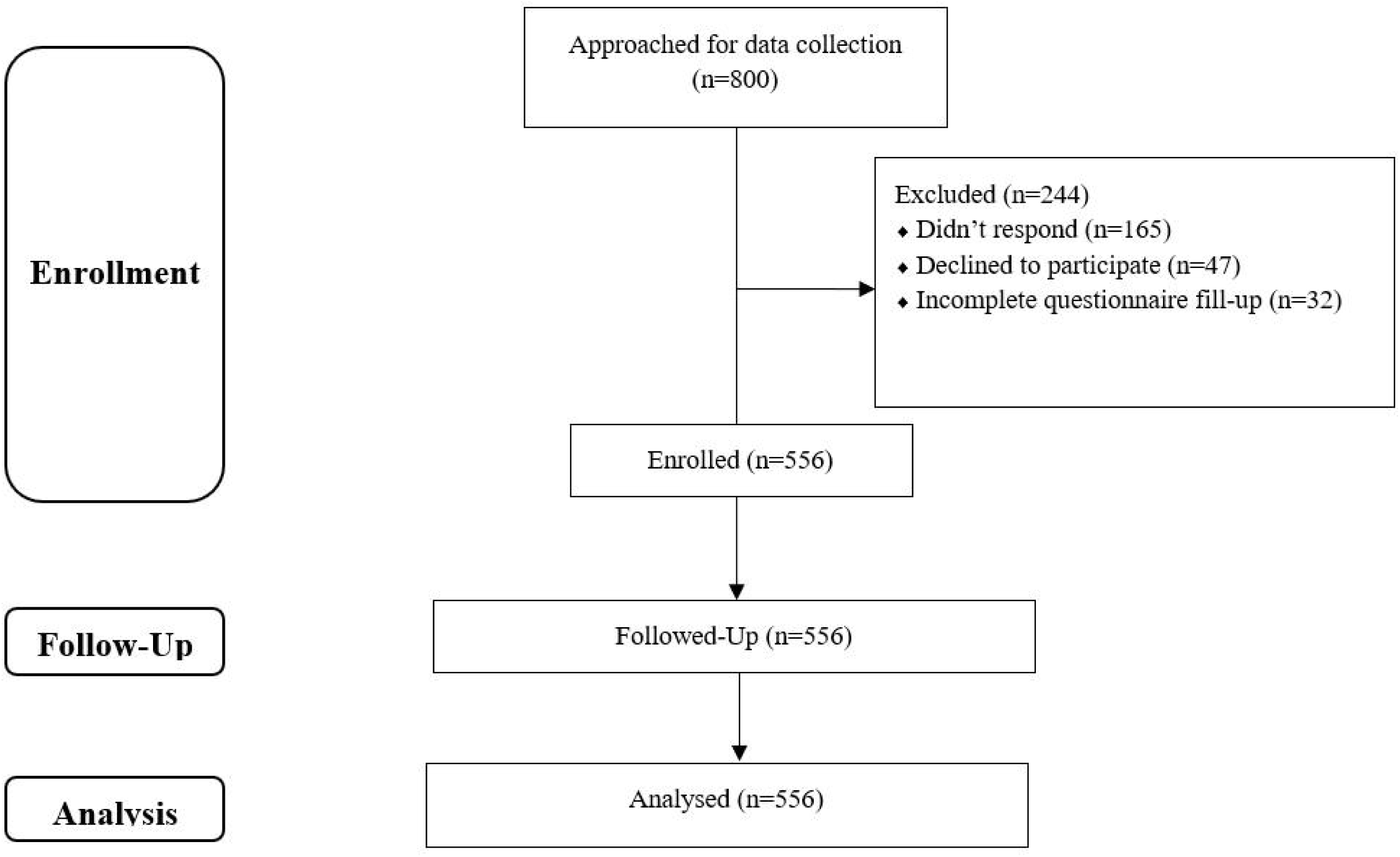

### Instrument and Measurement

A semi-structured and self-reported questionnaire including informed consent and four categories (sociodemographic, knowledge, attitudes, and perceptions) was used during data collection. The entire questionnaire was inputted into Google Forms without any randomization of items for online distribution and tested for usability and technical functionality. The form had 55 questions distributed over two pages. Mandatory items were highlighted with a red asterisk and relevant non-response option was present. Respondents were able to review their answers through the back button and change their response if they deemed necessary. The survey never displayed a second time once the user had filled it in to prevent duplicate entries.

### Sociodemographic information

The questionnaire included age, sex, marital status, educational institution, employment, monthly family income, and religion. In terms of personal information, individuals were queried about their medical history and sexual exposure.

### Knowledge, attitudes, and perceptions

To ascertain respondents’ level of knowledge, attitudes, and perceptions, a total of 46 structured questions were used (34-items for knowledge, 5-items for attitudes, and 7-items for perceptions). These questions were derived from a review of the prior literature.

The knowledge portion had 34 questions with three response categories (i.e., “True,” “False,” and “Don’t know”). These questions aimed to explore general knowledge, disease-specific knowledge, drug-specific knowledge, investigation-specific knowledge, healthy practice-specific knowledge, and knowledge about the associated factor. The ‘Correct answer’ was assigned a value of 1, while the ‘Wrong answer/Don’t know’ was assigned a 0. The total score was calculated by adding the raw scores for all 34 questions, ranging from 0 to 34. A score more than the mean suggested “Good knowledge” about male infertility. Cronbach’s alpha value for knowledge items was 0.861.

The attitude part included five statements (e.g., Mass media should promote educational programs on male infertility; Campaigns on Reproductive Health should be organized in the educational institutions), with a two-point Likert scale (0 = Disagree, 1 = Agree). The total score was determined by adding the raw values for each of the five statements, ranging from 0 to 5, and a score greater than the mean indicating a “Positive attitude” regarding male infertility. Cronbach’s alpha coefficient for attitudes items was 0.77.

The perceptions section included seven items assessing participants’ attitudes toward male infertility, six of which were yes/no questions (e.g., General people have poor knowledge about male infertility; Men feel depressed to be considered themselves as infertile etc.) and one were scenario-based questions (e.g., Suppose you are treating a couple for infertility, who should be investigated first?).

### Survey Administration

Trained research assistants contacted prospective participants via convenience and quota sampling and described the research in detail. Once the individuals were ascertained of meeting the inclusion criteria and consented to voluntary participation in the study, link to a web-based survey created by Google Forms was sent via facebook message/email/SMS making it a closed survey. The survey wasn’t announced or advertised anywhere else. Of the 588 eligible participants who agreed to participate 556 participants completed the entire questionnaire (completion rate: 94.56%), incomplete questionnaires were excluded from analysis.

### Statistical analysis

We used Stata (version 16; StataCorp, College Station, TX, USA) for data analysis. A histogram, a normal Q-Q plot, and the Kolmogorov-Smirnov test were used to check for normality in continuous data. Arithmetic mean was used for quantitative data as a measure of center and standard deviation was used as measure of dispersion. Chi-square tests and two-independent sample t-test were used to examine the relation of the mean knowledge and attitude scores with sociodemographic characteristics. Finally, we performed binary logistic regression analysis to find out the predictors of good knowledge and positive attitude. All p-values were considered statistically significant if <□0.05. Cronbach’s alpha was used to assess internal consistency for each knowledge and attitude scale.

### Ethics

The Institutional Review Board of North South University approved the research, and all participants provided informed consent. Wherever feasible, the 1964 Declaration of Helsinki and later modifications and comparable ethical standards were followed. Data collection was voluntary and no incentives was offered to participants. Data were only accessible to the authors and was not disclosed anywhere. All the reporting was done according to the Checklist for Reporting Results of Internet E-Surveys (CHERRIES) guidelines.^11^

## Result

### Sociodemographic Characteristics

Among the 556 participants, the majority (41.1%) was between 23 and 26 years of age, with females representing 59.89% of the respondents. 78.6% of the participants were Muslims, and 70.86% of the study participants were unmarried. Approximately 14.21% of participants had children, 10.07% of the participants were married but not planning for children currently, 2.16% were trying to conceive, 10.07% were not planning for children, and 1.44% were suffering from infertility. Among the respondents, 75.36% were from government institutions, 21.94% were from private institutions, and 2.7% were in the autonomous category. 37.77% of the participants were doctors, whereas nurses made up 10.61%, students were 44.24%, and others were 7.37%. About 71.22% of the participants had never been sexually exposed, but 28.78% gave a history of sexual exposure. The study found that 91.73 % of participants had no family history of infertility, while 8.27% had family history of infertility. 10.25% reported obesity as a co-morbidity, 7.37% reported hypertension, 5.22% reported diabetes, 3.06% reported a history of mumps, 3.06% reported a history of genitourinary infection, 1.62% reported autoimmune diseases, 1.26% reported a history of tuberculosis, and 76.44% reported no personal history of diseases. In terms of household income, details of the participants have been mentioned in **Table 1**.

**Table 1:**
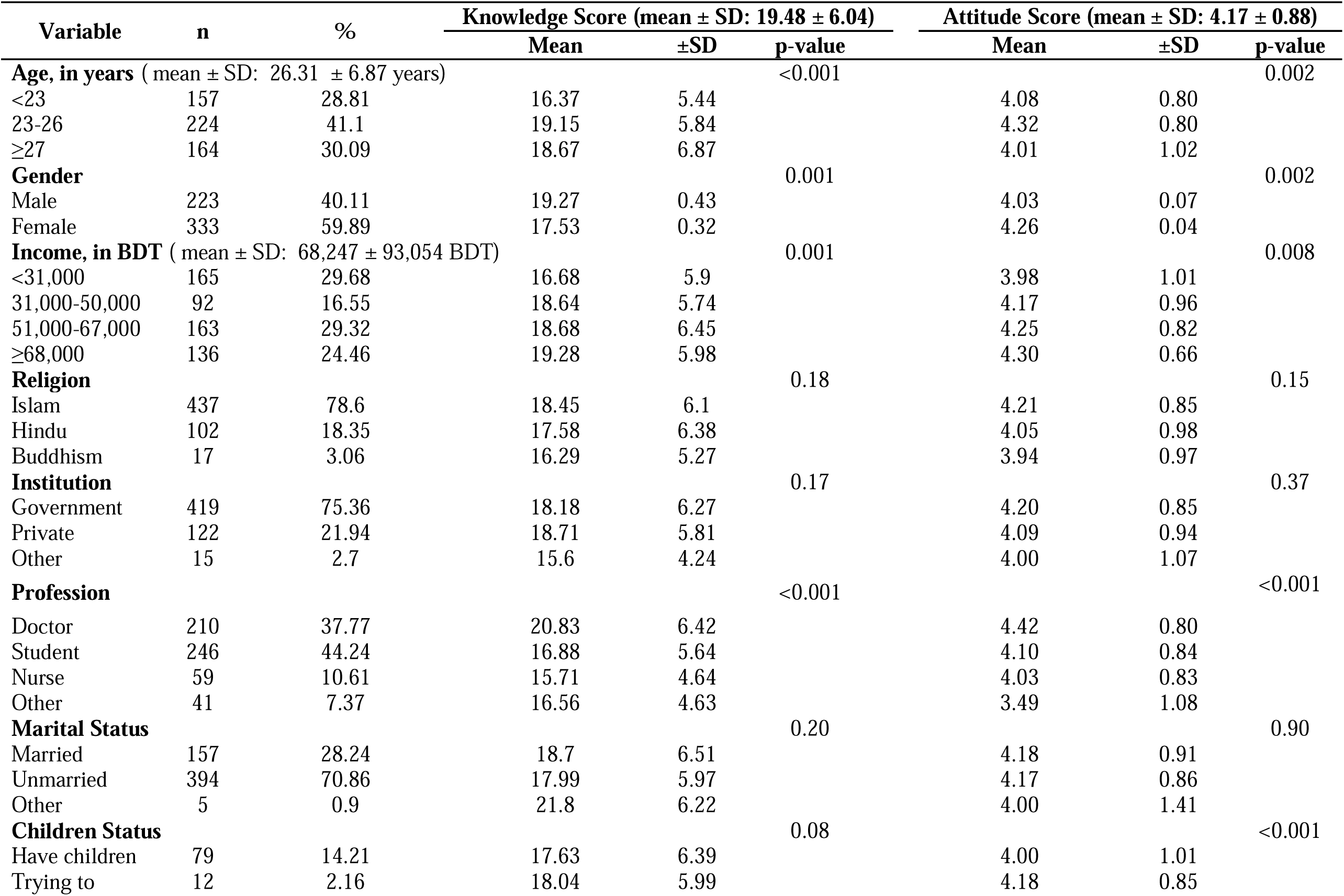
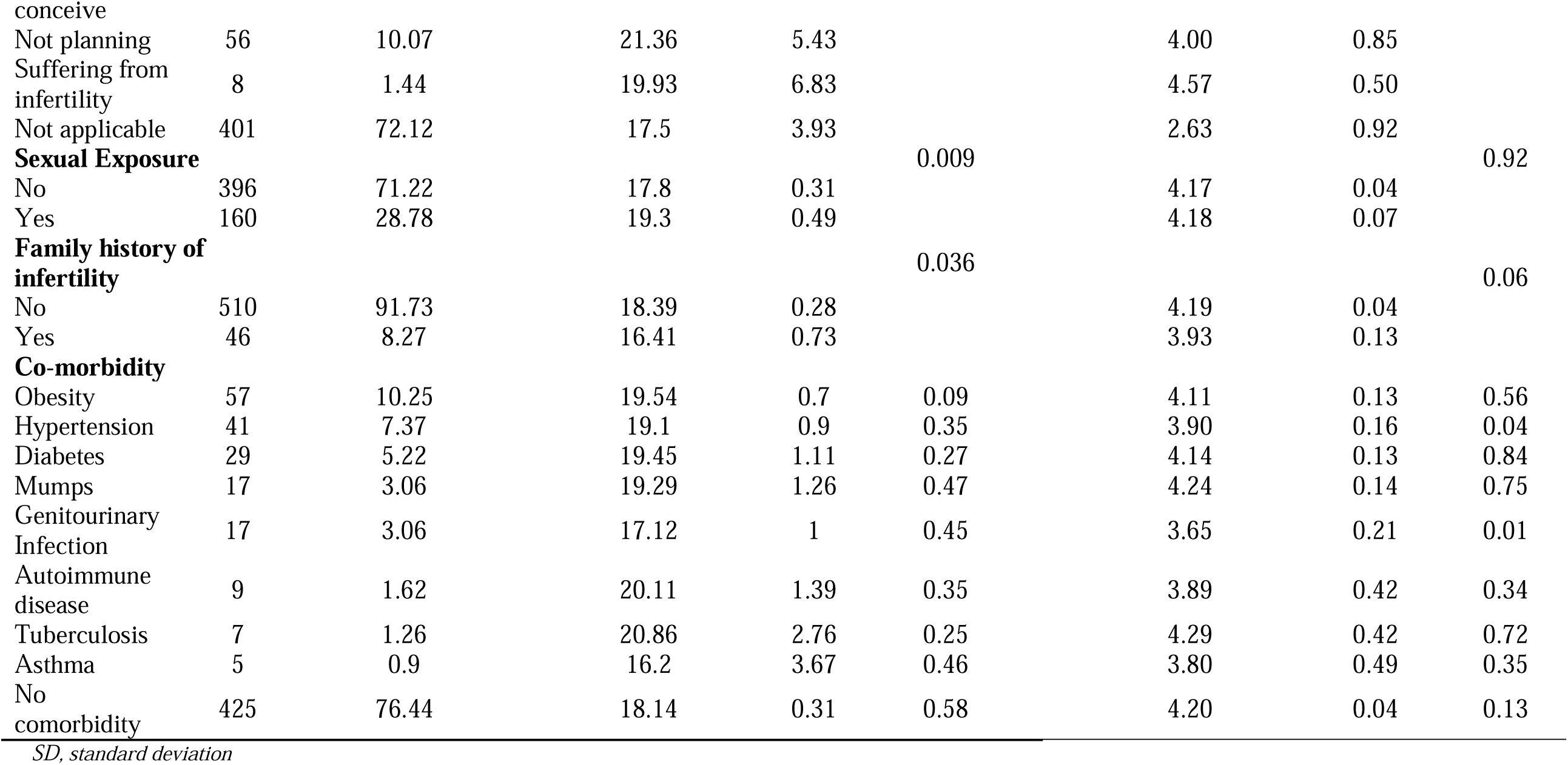
Baseline characteristics of study participants and their knowledge & attitude score (n=556)

### Knowledge about Male Infertility

In this section, the maximum possible score for each respondent was 34, whereas the respondents’ mean score was 19.48 ± 6.04. The overall prevalence of good knowledge was 50.18 %. The highest overall prevalence of good knowledge was regarding investigation-related information (83.09%), whereas the lowest overall prevalence was in the disease-related section (38.74%). More than half of the males (56.5%) showed good knowledge, although for females was 45.05%. Concerning different subgroups, 74.03% of MBBS doctors showed good knowledge; on the other hand, only 28.81% of nurses showed good knowledge. Among the other subgroups, 57.14% BDS Doctors, 42.02% MBBS Students, 41.38% BDS Students, and 31.71% other healthcare staff had overall good knowledge.

### Attitude towards Male Infertility

The overall total possible score for attitude for each respondent was 5, but the mean score of the respondents was mean: 4.17 ± 0.88. Surprisingly only 39.21% had a positive attitude towards male infertility. The most positive attitude was regarding ‘Mass media should promote educational programs on male infertility, where around 96% of respondents had a positive attitude. On the other hand, only 46% of the respondents had a positive attitude regarding the treatment cost of male infertility **(Table 4)**.

### Perception Regarding Male Infertility

93.71 % participants perceived, “general people have poor knowledge about male infertility” and 75.72 % thinks, “Uses of alternative medicine (herbal, spiritual healing, homeopathy etc.) are popular in male infertility treatment.” Concurrently, 89.39 % believe that males are depressed about their infertility. Approximately 94.96 % believe that males who are infertile endure a challenging emotional journey. Only 7.19% of the participants took part in male infertility training. About 84% disagreed with the notion that “prescription of alternative medicine is beneficial in the treatment of male infertility.” During infertility treatment, 14.57 % believed “male should be investigated first,” 8.63 % thought “female should be investigated first,” and 76.8 % considered “both male and female should be investigated first.”

### Predictors of Knowledge and Attitude towards Male Infertility

**Table 1** presents the bivariate analysis of potential factors associated with the knowledge and attitude towards male infertility. Two independent sample t-test revealed age, gender, income, profession, and children status were significantly associated with both knowledge and attitude. In addition, sexual exposure was significantly associated with knowledge but not with attitude. Family history of infertility was also significantly associated with both knowledge and attitude. In the case of co-morbidities, obesity was significantly associated with knowledge of male infertility. On the other hand, hypertension and a history of genitourinary infection were significantly associated with attitudes towards male infertility.

**Table 2** reveals that males have significantly better knowledge than females in general, drug-related and associated factors related knowledge items. MBBS doctors had significantly better knowledge than all others professions in all the knowledge domains except associated factors related knowledge, and they were significantly better than BDS doctors in general, disease-related and drug-related items.

**Table 2:**
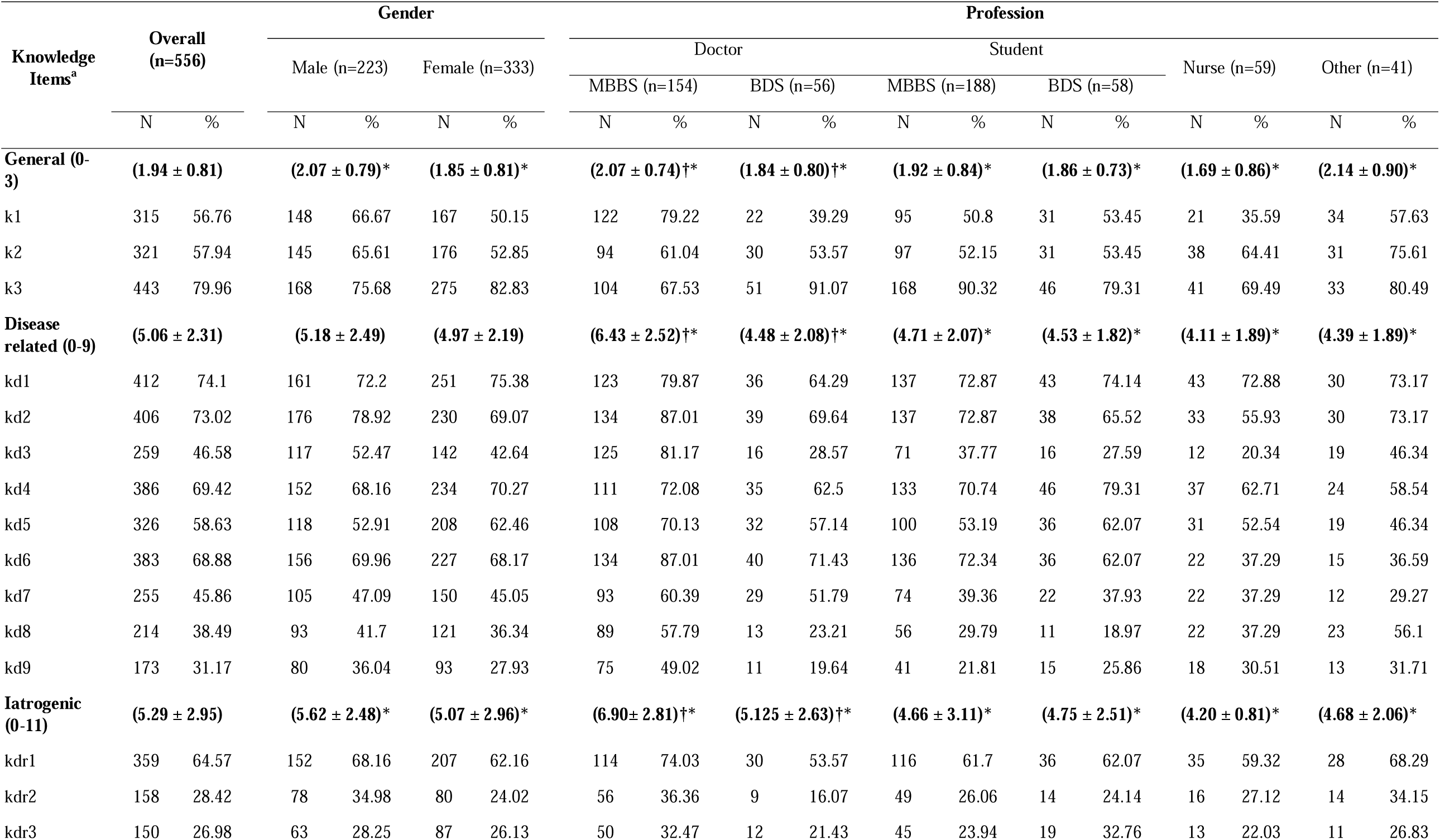
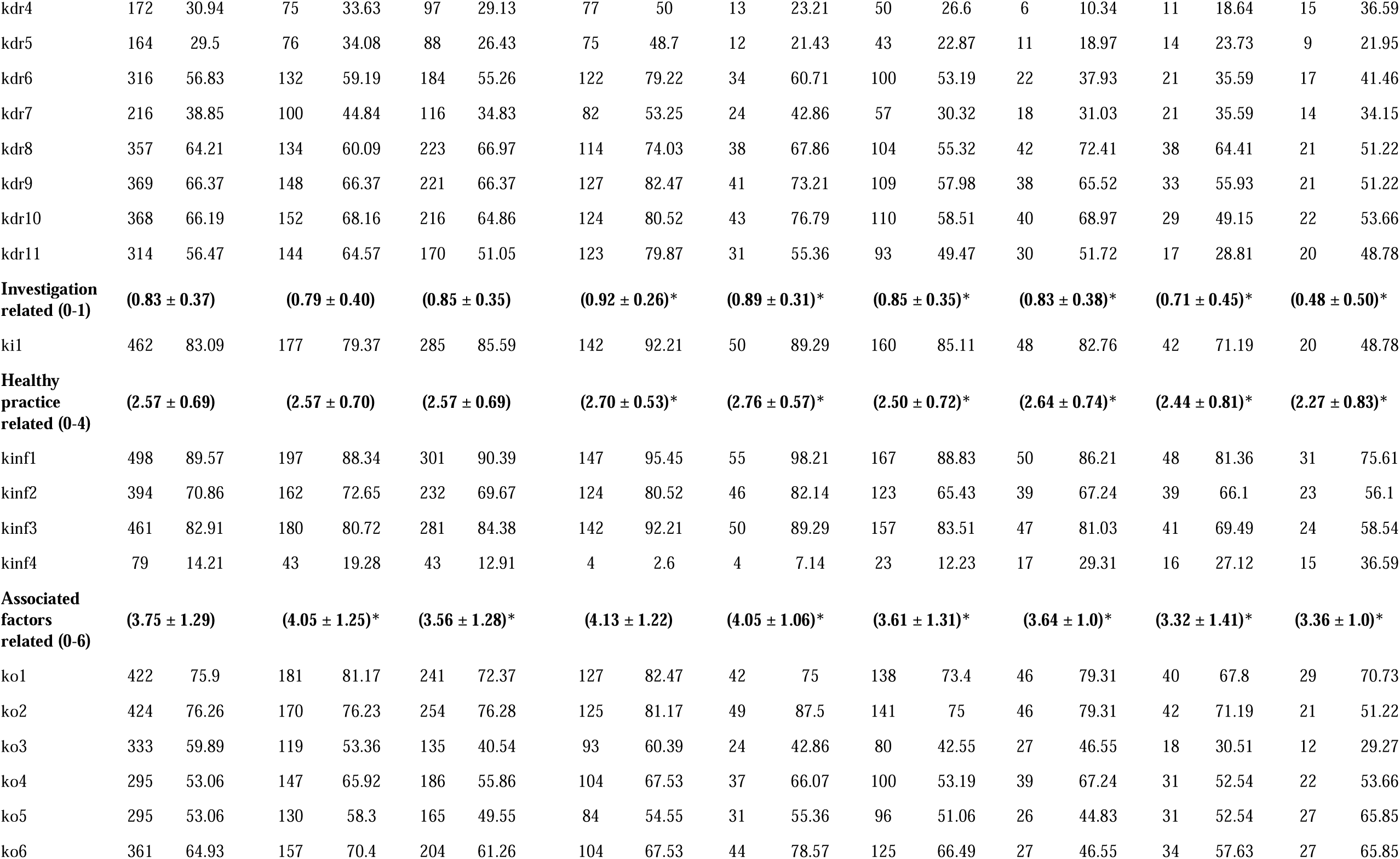
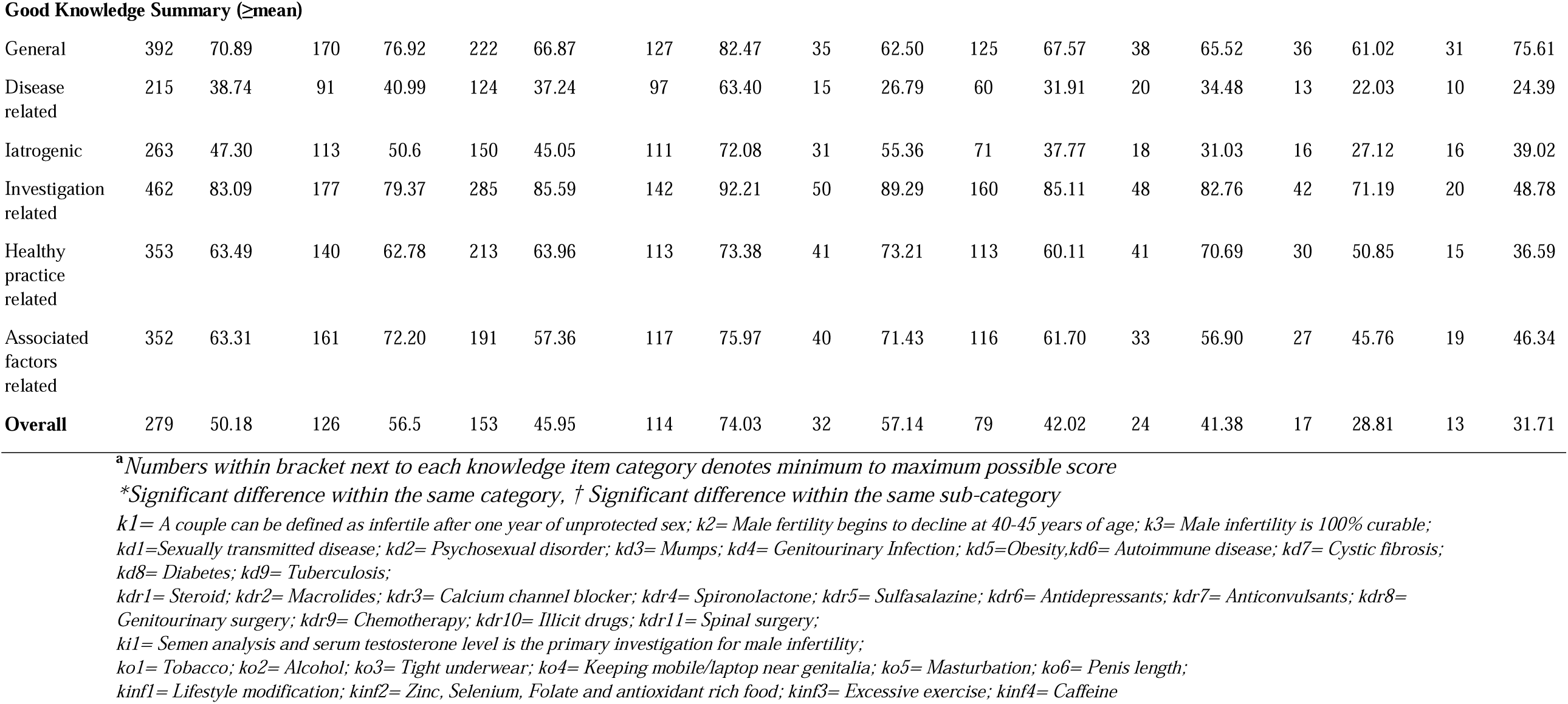
Correct knowledge regarding male infertility in gender and profession subgroups (n=556)

**Table 3** and **Table 5** depict the binary logistic regression model for good knowledge and positive attitude, respectively. Participants between 23 and 26 had 80% more knowledge than participants whose age was less than 23 years (OR: 1.81; 95% CI: 1.099 to 2.988). In addition, participants from private institutions have 9% more knowledge about male infertility than those from government institutions (OR: 1.099; 95% CI: 0.69 to 1.755). MBBS doctors had the most knowledge among all the professions. Compared to them, BDS doctors, MBBS students, BDS students, nurses, and other health professionals had 51%, 65%, 66%, 83%, and 74% less knowledge about male infertility. Furthermore, participants with a positive attitude towards male infertility had around 60% more knowledge than participants with negative attitudes (OR: 1.59; 95% CI: 1.082 to 2.342).

**Table 3:** Binary logistic regression analysis of factors affecting good knowledge of respondents regarding male infertility (n=556)

| Variable | AOR | p-value | 95% CI |  |  |
| --- | --- | --- | --- | --- | --- |
| **Age (year)** |  |  |  |  |  |
| <23 | ref. |  |  |  |  |
| 23-26 | 1.812 | 0.020 | 1.099 | to | 2.988 |
| ≥27 | 1.546 | 0.186 | 0.811 | to | 2.949 |
| **Institution** |  |  |  |  |  |
| Government | ref. |  |  |  |  |
| Private | 1.099 | 0.692 | 0.688 | to | 1.755 |
| Other | 0.141 | 0.017 | 0.029 | to | 0.702 |
| **Profession** |  |  |  |  |  |
| MBBS Doctor | ref. |  |  |  |  |
| BDS Doctor | 0.499 | 0.043 | 0.254 | to | 0.977 |
| MBBS Student | 0.356 | 0.001 | 0.198 | to | 0.638 |
| BDS Student | 0.342 | 0.003 | 0.167 | to | 0.702 |
| Nurse | 0.174 | 0.000 | 0.086 | to | 0.350 |
| Other | 0.260 | 0.002 | 0.112 | to | 0.602 |
| **Attitude** |  |  |  |  |  |
| Negative Attitude | ref. |  |  |  |  |
| Positive Attitude | 1.592 | 0.018 | 1.082 | to | 2.342 |

*AOR, adjusted odds ratio; CI, confidence interval*

**Table 4:**
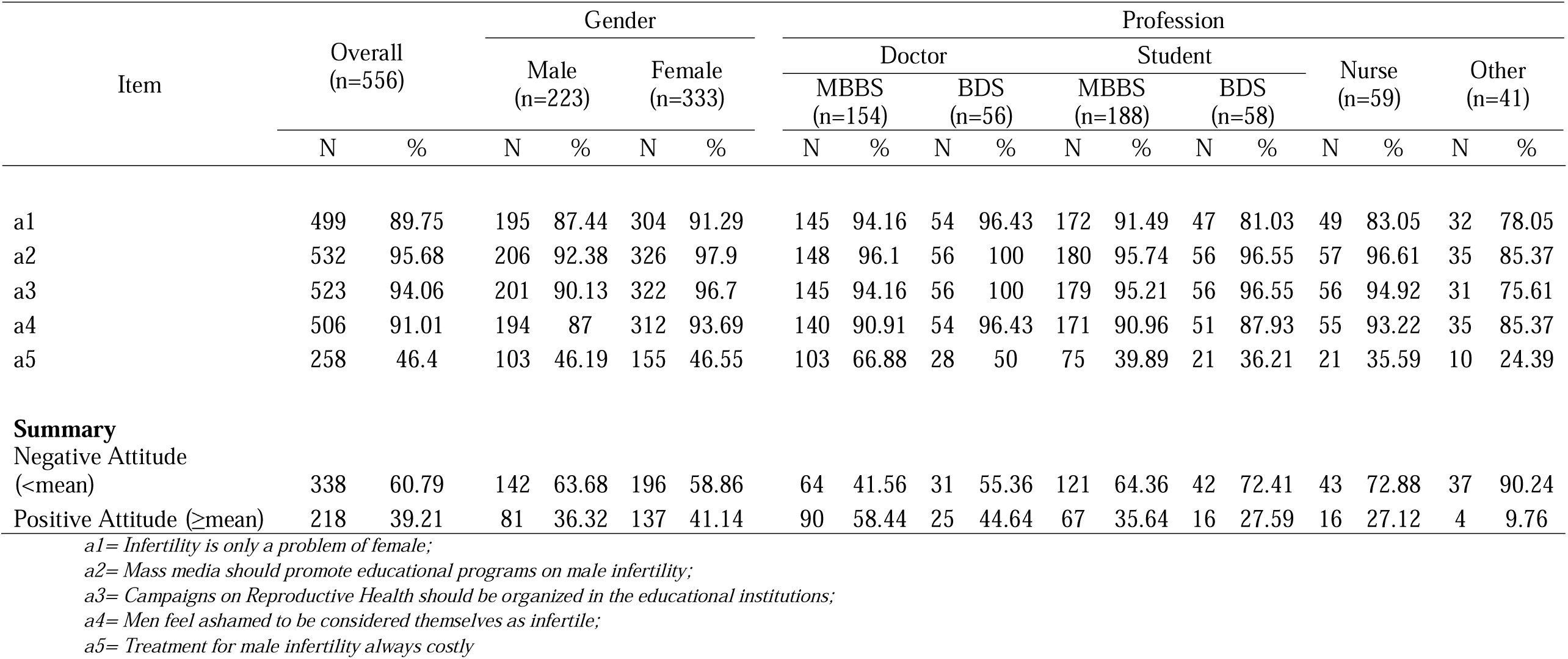
Attitude regarding male infertility results in gender and profession subgroups (n=556)

| Item | Overall<br>(n=556) |  | Gender |  |  |  | Profession |  |  |  |  |  |  |  |  |  |  |  |
| --- | --- | --- | --- | --- | --- | --- | --- | --- | --- | --- | --- | --- | --- | --- | --- | --- | --- | --- |
|  |  |  | Male<br>(n=223) |  | Female<br>(n=333) |  | Doctor |  |  |  | Student |  |  |  | Nurse<br>(n=59) |  | Other<br>(n=41) |  |
|  |  |  |  |  |  |  | MBBS<br>(n=154) |  | BDS<br>(n=56) |  | MBBS<br>(n=188) |  | BDS<br>(n=58) |  |  |  |  |  |
|  | N | % | N | % | N | % | N | % | N | % | N | % | N | % | N | % | N | % |
| a1 | 499 | 89.75 | 195 | 87.44 | 304 | 91.29 | 145 | 94.16 | 54 | 96.43 | 172 | 91.49 | 47 | 81.03 | 49 | 83.05 | 32 | 78.05 |
| a2 | 532 | 95.68 | 206 | 92.38 | 326 | 97.9 | 148 | 96.1 | 56 | 100 | 180 | 95.74 | 56 | 96.55 | 57 | 96.61 | 35 | 85.37 |
| a3 | 523 | 94.06 | 201 | 90.13 | 322 | 96.7 | 145 | 94.16 | 56 | 100 | 179 | 95.21 | 56 | 96.55 | 56 | 94.92 | 31 | 75.61 |
| a4 | 506 | 91.01 | 194 | 87 | 312 | 93.69 | 140 | 90.91 | 54 | 96.43 | 171 | 90.96 | 51 | 87.93 | 55 | 93.22 | 35 | 85.37 |
| a5 | 258 | 46.4 | 103 | 46.19 | 155 | 46.55 | 103 | 66.88 | 28 | 50 | 75 | 39.89 | 21 | 36.21 | 21 | 35.59 | 10 | 24.39 |
| Summary |  |  |  |  |  |  |  |  |  |  |  |  |  |  |  |  |  |  |
| Negative Attitude (<mean) | 338 | 60.79 | 142 | 63.68 | 196 | 58.86 | 64 | 41.56 | 31 | 55.36 | 121 | 64.36 | 42 | 72.41 | 43 | 72.88 | 37 | 90.24 |
| Positive Attitude (≥mean) | 218 | 39.21 | 81 | 36.32 | 137 | 41.14 | 90 | 58.44 | 25 | 44.64 | 67 | 35.64 | 16 | 27.59 | 16 | 27.12 | 4 | 9.76 |

a1= Infertility is only a problem of female;
a2= Mass media should promote educational programs on male infertility;
a3= Campaigns on Reproductive Health should be organized in the educational institutions;
a4= Men feel ashamed to be considered themselves as infertile;
a5= Treatment for male infertility always costly

**Table 5:** Binary logistic regression analysis of factors affecting the positive attitude of respondents regarding male infertility (n=556)

|  | AOR | p-value | 95% CI |  |  |
| --- | --- | --- | --- | --- | --- |
| Gender |  |  |  |  |  |
| Male | ref. |  |  |  |  |
| Female | 1.483 | 0.049 | 1.002 | to | 2.194 |
| Profession |  |  |  |  |  |
| MBBS Doctor | ref. |  |  |  |  |
| BDS Doctor | 0.560 | 0.073 | 0.298 | to | 1.055 |
| MBBS Student | 0.409 | 0.000 | 0.256 | to | 0.652 |
| BDS Student | 0.289 | 0.000 | 0.147 | to | 0.568 |
| Nurse | 0.272 | 0.000 | 0.135 | to | 0.548 |
| Other | 0.097 | 0.000 | 0.033 | to | 0.289 |
| Knowledge |  |  |  |  |  |
| Poor Knowledge | ref. |  |  |  |  |
| Good Knowledge | 1.610 | 0.013 | 1.105 | to | 2.346 |

*AOR, adjusted odds ratio; CI, confidence interval*

**Table 6:** Perceptions of the respondents regarding male infertility (n=556)

| Questions pertaining to perception and practice | Response | Total |  |
| --- | --- | --- | --- |
|  |  | n | % |
| General population have poor knowledge about male infertility | No | 35 | 6.29 |
|  | Yes | 521 | 93.71 |
| Uses of alternative medicine (herbal, spiritual healing, homeopathy etc.) are popular in male infertility treatment. | No | 135 | 24.28 |
|  | Yes | 421 | 75.72 |
| Men feel depressed to be considered themselves as infertile. | No | 59 | 10.61 |
|  | Yes | 497 | 89.39 |
| Many men diagnosed with male infertility, face a difficult emotional journey- do you agree or not? | No | 28 | 5.04 |
|  | Yes | 528 | 94.96 |
| Did you participate in any training on male infertility? | No | 516 | 92.81 |
|  | Yes | 40 | 7.19 |
| Do you support prescribing alternative medicines (herbal, spiritual healing etc.) are helpful in the treatment of male infertility? | No | 462 | 83.09 |
|  | Yes | 94 | 16.91 |
| Suppose you're treating a couple for infertility, who should be investigated first? | Male | 81 | 14.57 |
|  | Female | 48 | 8.63 |
|  | Both | 427 | 76.8 |

In terms of attitude, **Table 5** shows that females had a 48% more positive attitude towards male infertility than males, with a 95% confidence interval (OR=1.48; 95%CI: 1.002 and 2.19, p=0.049). In addition, BDS doctors, MBBS students, BDS students, nurses, and other health professionals had 44%, 60%, 72%, 73%, and 91% less positive attitudes towards male infertility than MBBS doctors. Moreover, participants with good knowledge about male infertility had 61% more positive attitudes compared to participants who had poor knowledge (OR=1.61; 95% CI: 1.105 and 2.346, p=0.013).

## Discussion

This study explored the knowledge, attitude and beliefs of medical students and healthcare professionals of Bangladesh regarding male infertility. The key findings were that nearly half of the study population didn’t have a good male infertility knowledge and nearly two-thirds had negative attitudes regarding male infertility. Young (23-26 years) healthcare professionals and medical students were more likely to have good knowledge. Surprisingly, females were more likely to have positive attitude compared to males. Among all the professions, MBBS doctors were most likely to have good knowledge and positive attitude regarding male infertility. Good knowledge of male infertility predicted positive attitude and vice versa.

This study clearly highlighted the lack of awareness concerning male infertility as barely half (50.18%) of the population in our study possessed overall good knowledge. Knowledge is also significantly (p=0.001) lower among females compared to males. This lack of understanding explains why infertility is associated with such a negative connotation in our society.^12^ Our finding is consistent with a global poll of nearly 17,500 women from ten countries, conducted during the Word Fertility Awareness Month (2006).^13^ The lack of awareness was further reinforced when it was discovered that just half (56.76%) of the participants recognized how infertility is diagnosed after at least one year of regular unprotected sex. This may influence the couple’s decision to seek treatment, which should not be premature or postponed.^14^ Most of our participants were aware of the age at which male fertility begins to diminish which corroborated the findings of Hammarberg et al (2013).^15^ Despite advances in microsurgery and genetics that have transformed diagnosis and treatment of male infertility, there are still many unanswered questions, as properly identified by most of our participants (79.96%).^16^

A number of factors have been associated with male infertility; including various diseases (e.g., STD etc.), treatment modalities (e.g., steroid etc.) and lifestyle habits (e.g., smoking, alcohol consumption etc.).^17–19^ While it is unnecessary for non-physicians to be aware of all possible causes, it is necessary for them to be aware of acquired and potentially preventable causes of infertility, such as sexually transmitted illnesses. In contrast to Ali et al. (2011) majority of our participants didn’t have a good knowledge regarding disease related (38.74%) and iatrogenic causes (47.30%) of male infertility.^20^ However, most of them correctly identified STDs, psychosexual disorders, genitourinary infections, obesity and autoimmune diseases to be the causes of male infertility.

Nearly three-fourth of our participants accurately identified the negative consequences of cigarette smoking and alcohol consumption on male fertility. These findings agree with data from previous studies.^20,21^ This higher prevalence of awareness regarding tobacco and alcohol may be due to extensive campaigning by the Government of Bangladesh and also associated social stigma.^22,23^

People all over the world have accumulated a plethora of myths about reproductive health and fertility. We found several misunderstandings regarding of male infertility in our study, such as keeping mobile near genitalia (46.94%), masturbation (46.94%), wearing tight underwear (40.11%), length of penis (35.07%) & excessive exercise (17.09%). These beliefs are consistent with studies from Pakistan and Saudi Arabia.^20,24^

Age of the participants was a significant predictor of good knowledge; individuals from 23 to 26 years of age having the maximum likelihood (AOR=1.812, p=0.02) of having good knowledge regarding male infertility. Hammarberg et al. (2013) came to similar conclusion from their study on Australian population of reproductive age.^15^

Females in our study were 48.3% more prone to having a positive attitude towards male infertility compared to males (AOR=1.483, p=0.049). Women who are infertile are often subjected to more societal pressure than men, which may explain why women a more positive attitude on infertility than men.^25^

MBBS doctors had a significantly higher prevalence of good knowledge and positive attitude compared to all other healthcare professionals and medical students, this may be explained by their medical background and training. Zhang et al. (2019) reports similar positive attitudes but limited knowledge among oncology physicians of China.^26^

93.71% of our study participants perceive general population to have poor knowledge about male infertility, which in fact corresponds to the finding of our current study. This poor knowledge may be attributed to only 7.19% receiving training on male infertility. When asked who should be investigated first in case of an infertile couple, majority (76.8%) of the participants responded “both” which is consistent with the findings from Saudi Arabia but contrasts findings from Latin America.^24,27^ Nearly all of the study participants perceive men diagnosed with male infertility face a difficult emotional journey (94.96%) and often feel depressed about their situation (89.39%) which is in line with the current evidence.^28^ Although 83.09% don’t support alternative medicine (homeopathy, spiritual healing etc.) for the treatment of male infertility, majority (75.72%) admits these treatment modalities are popular in the treatment of male infertility. This shows how common it is for people to think that all illnesses can’t be cured by medical science and this belief is congruent to the evidence from Pakistan.^20^

## Limitations

Owing to our study’s cross-sectional design, we cannot infer causality for the associations that we have reported in this article. We, therefore, sought to control for the potential effect of confounders by reporting AORs using multiple regression models. Number of participants in our study may be a limitation, although it is larger than that of similar studies.^15,20,21^ Furthermore, selection bias is always a potential drawback in non-random studies. We employed quota sampling technique to ensure representative sample from all over the country.

## Conclusion

Aside from these caveats, the findings of our current study suggest that healthcare professionals and medical students of Bangladesh have overall inadequate knowledge and negative attitude towards male infertility. This points to the necessity for multidisciplinary training programs, the formulation of a standard guideline for healthcare workers, and necessary curriculum changes for medical students.

## Data Availability

All data produced in the present work are contained in the manuscript

## References

1. Kshrisagar SP, Shirsath AS. A cross-sectional study of fertile period awareness, knowledge, attitudes and practice in infertile couples seeking fertility assistance. International Journal of Reproduction, Contraception, Obstetrics and Gynecology. 2018;7(9):3744. doi:10.18203/2320-1770.ijrcog20183787

2. Ombelet W, Cooke I, Dyer S, Serour G, Devroey P. Infertility and the provision of infertility medical services in developing countries. Human reproduction update. 2008;14(6):605–621. doi:10.1093/humupd/dmn042

3. Kshrisagar SP, Shirsath AS. A cross-sectional study of fertile period awareness, knowledge, attitudes and practice in infertile couples seeking fertility assistance. International Journal of Reproduction, Contraception, Obstetrics and Gynecology. 2018;7(9):3744. doi:10.18203/2320-1770.ijrcog20183787

4. Rahman F, Rahman M, Mahmud N, Ahsan GU, Islam MI. Prevalence of Male Infertility among the Infertile Couples Attended at BIRDEM General Hospital, Dhaka. Ibrahim Cardiac Medical Journal. 2018;6(1-2):25–32. doi:10.3329/icmj.v6i1-2.53754

5. Chowdhury SH. Male Infertility-A Global Overview & Bangladesh Perspective. Journal of Current Medical Practice. Published online 2019:32.

6. Kamala E, Karunya AS, Nalinakumari SD. Knowledge, Attitude and Practice about Male Infertility among Men and Women in the Field Practice Area of a Tertiary Care Teaching Hospital. INTERNATIONAL JOURNAL OF PREVENTIVE AND PUBLIC HEALTH SCIENCES. 2019;5(2):1–4.

7. Daumler D, Chan P, Lo KC, Takefman J, Zelkowitz P. Men’s knowledge of their own fertility: a population-based survey examining the awareness of factors that are associated with male infertility. Human Reproduction. 2016;31(12):2781–2790. doi:10.1093/humrep/dew265

8. Abolfotouh MA, Alabdrabalnabi AA, Albacker RB, Al-Jughaiman UA, Hassan SN. Knowledge, attitude, and practices of infertility among Saudi couples. International journal of general medicine. 2013;6:563–573. doi:10.2147/IJGM.S46884

9. Fahami F, Quchani SH, Ehsanpour S, Boroujeni AZ. Lived experience of infertile men with male infertility cause. Iranian journal of nursing and midwifery research. 2010;15(Suppl 1):265-271.

10. Joja OD, Dinu D, Paun D. Psychological Aspects of Male Infertility. An Overview. Procedia - Social and Behavioral Sciences. 2015;187:359–363. doi:https://doi.org/10.1016/j.sbspro.2015.03.067

11. Eysenbach G. Improving the quality of Web surveys: the Checklist for Reporting Results of Internet E-Surveys (CHERRIES). Journal of medical Internet research. 2004;6(3). doi:10.2196/JMIR.6.3.E34

12. Papreen N, Sharma A, Sabin K, Begum L, Ahsan SK, Baqui AH. Living with infertility: Experiences among urban slum populations in Bangladesh. Reproductive Health Matters. 2000;8(15):33–44. doi:10.1016/S0968-8080(00)90004-1

13. World Fertility Awareness Month 2006. What You Never Know about Fertility..; 2006.

14. Crosignani PG, Albertini DF, Anderson R, et al. A prognosis-based approach to infertility: understanding the role of time. Human Reproduction. 2017;32(8):1556–1559. doi:10.1093/HUMREP/DEX214

15. Hammarberg K, Setter T, Norman RJ, Holden CA, Michelmore JE, Johnson LE. Knowledge about factors that influence fertility among Australians of reproductive age: a population-based survey. Fertility and Sterility. 2013;99:502–507. doi:10.1016/j.fertnstert.2012.10.031

16. Wald M. Male infertility: Causes and cures. Sexuality, Reproduction and Menopause. 2005;3(2):83–87. doi:10.1016/J.SRAM.2005.09.006

17. Fode M, Fusco F, Lipshultz L, Weidner W, Catto J. Sexually Transmitted Disease and Male Infertility: A Systematic Review. Published online 2016. doi:10.1016/j.euf.2016.08.002

18. Pasqualotto FF, Lucon AM, Sobreiro BP, Pasqualotto EB, Arap S. Review Effects of Medical Therapy, Alcohol, Smoking, and Endocrine Disruptors on Male Infertility.

19. el Osta R, Almont T, Diligent C, Hubert N, Eschwège P, Hubert J. Anabolic steroids abuse and male infertility. Published online 2016. doi:10.1186/s12610-016-0029-4

20. Ali S, Sophie R, Imam AM, et al. Knowledge, perceptions and myths regarding infertility among selected adult population in Pakistan: A cross-sectional study. BMC Public Health. 2011;11. doi:10.1186/1471-2458-11-760

21. Nakhon SN, Limvorapitux P, Vichinsartvichai P. Knowledge regarding factors that influence fertility in Thai reproductive-age population living in urban area: A cross-sectional study. Clinical and Experimental Reproductive Medicine. 2018;45(1):38–43. doi:10.5653/cerm.2018.45.1.38

22. Islam JY, Zaman MM, Bhuiyan MR, et al. Alcohol consumption among adults in Bangladesh: results from STEPS 2010. WHO South-East Asia Journal of Public Health. 2017;6(1).

23. National Tobacco Control Cell. Mass Media Campaign. Health Services Division, Ministry of Health & Family Welfare, Government of the People’s Republic of Bangladesh. Accessed February 15, 2022. https://ntcc.gov.bd/activities/mass-media-campaign

24. Abolfotouh MA, Alabdrabalnabi AA, Albacker RB, Al-Jughaiman UA, Hassan SN. Knowledge, attitude, and practices of infertility among Saudi couples. International Journal of General Medicine. 2013;6:563–573. doi:10.2147/IJGM.S46884

25. Bassey Iabasi E, Isiwele EM, Omotoso A, Ushie ED, Ekwere PD. Knowledge, Perceptions and Attitudes towards Male Infertility□: A Cross Sectional Survey in a Tertiary Institution in South-southern Nigeria. Ijdmsr. 2018;2(4):22–28.

26. Zhang H, Wang G, Jiang B, et al. The Knowledge, Attitude, and Self-Reported Behaviors of Oncology Physicians Regarding Fertility Preservation in Adult Cancer Patients. Journal of Cancer Education. 2020;35(6):1119–1127. doi:10.1007/s13187-019-01567-6

27. Luna F. Assisted reproductive technologies in Latin America: some ethical and sociocultural issues. In: Vayena E, Rowe P, Griffin P, eds. Current Practices and Controversies in Assisted Reproduction. World Health Organization; 2002:31–34. Accessed February 15, 2022. http://apps.who.int/iris/bitstream/handle/10665/42576/9241590300.pdf;jsessionid=45C4C8B5745FB772CF10405F3454FF55?sequence=1

28. Ahmadi H, Montaser-Kouhsari L, Nowroozi MR, Bazargan-Hejazi S. Male infertility and depression: a neglected problem in the Middle East. The journal of sexual medicine. 2011;8(3):824–830. doi:10.1111/J.1743-6109.2010.02155.X

